# Rifampicin mono-resistant tuberculosis is not the same as multidrug-resistant tuberculosis: a descriptive study from Khayelitsha, South Africa

**DOI:** 10.1101/2021.06.14.21258812

**Authors:** Zubeida Salaam-Dreyer, Elizabeth M. Streicher, Frederick A. Sirgel, Fabrizio Menardo, Sonia Borrell, Miriam Reinhard, Anna Doetsch, Patrick G.T. Cudahy, Erika Mohr-Holland, Johnny Daniels, Anzaan Dippenaar, Mark P. Nicol, Sebastien Gagneux, Robin M. Warren, Helen Cox

## Abstract

Rifampicin mono-resistant TB (RMR-TB) constitutes 38% of all rifampicin-resistant TB (RR-TB) in South Africa and is increasing. We aimed to compare RMR-TB with multidrug-resistant TB (MDR-TB) within a high TB, RR-TB and HIV burden setting. Patient-level clinical data and stored RR-TB isolates from 2008-2017 with available whole genome sequencing (WGS) data were used to describe risk factors associated with RMR-TB and to compare rifampicin-resistance (RR) conferring mutations between RMR-TB and MDR-TB. A subset of isolates with particular RR-conferring mutations were subjected to semi-quantitative rifampicin phenotypic drug susceptibility testing. Among 2,041 routinely diagnosed RR-TB patients, 463 (22.7%) had RMR-TB. HIV-positive individuals (adjusted Odds Ratio 1.4, 95% CI 1.1-1.9) and diagnosis between 2013-2017 versus 2008-2012 (aOR 1.3, 1.1-1.7) were associated with RMR-TB. Among 1,119 (54.8%) patients with available WGS data showing RR-TB, significant differences in the distribution of *rpoB* RR-conferring mutations between RMR-TB and MDR-TB isolates were observed. Mutations associated with high-level RR were more commonly found among MDR-TB isolates (811/889, 90.2% versus 162/230, 70.4% among RMR-TB, p<0.01). In particular, the *rpoB* L430P mutation, conferring low-level RR, was identified in 32/230 (13.9%) RMR-TB versus 10/889 (1.1%) in MDR-TB (p<0.01). Among 10 isolates with an *rpoB* L430P mutation, 7 were phenotypically susceptible using the critical concentration of 0.5 µg/ml (range 0.125-1 µg/ml). The majority (215/230, 93.5%) of RMR-TB isolates showed susceptibility to all other TB drugs, highlighting the potential benefits of WGS for simplified treatment. These data suggest that the evolution of RMR-TB differs from MDR-TB with a potential contribution from HIV infection.

## Introduction

Globally, an estimated 465,000 individuals became ill with rifampicin-resistant tuberculosis (RR-TB) in 2019.[1] Among these, 78% were estimated to have multidrug-resistant tuberculosis (MDR-TB) with resistance to both rifampicin (RIF) and isoniazid (INH), whilst the remainder had rifampicin mono-resistant TB (RMR-TB, RIF resistance and INH susceptibility). While RMR-TB represents 22% of all RR-TB globally, this percentage varies widely across high RR-TB burden countries, ranging from <1% in several countries to more than 40% in countries as diverse as Kenya and Tajikistan.[1] In South Africa, RMR-TB constitutes 38% of the more than 13,000 RR-TB cases diagnosed annually.[1] In addition, national TB drug resistance surveys have suggested that RMR-TB increased significantly between 2002 and 2012 in South Africa, while the proportion of all TB cases with MDR-TB remained relatively constant.[2]

RIF resistance in *Mycobacterium tuberculosis (M*.*tb)* is caused by mutations predominantly in the rifampicin-resistance determining region (RRDR) of the RNA polymerase β subunit (*rpoB*) gene.[3] While any non-synonymous mutation in the RRDR region is considered to confer RR, there is now increasing evidence that some *rpoB* mutations, often described as ‘disputed’ or ‘discordant’, are associated with decreased RIF susceptibility. The elevated minimum inhibitory concentrations (MICs) caused by these mutations show a range of values around both the epidemiological cut-off value and the critical concentration (CC).[4, 5] Associations between these low-level RIF resistant variants and poor patient outcomes[5-8] have led to a recent change in the CC value recommended by the World Health Organization (WHO) for RIF from 1.0 to 0.5 µg/ml in MGIT 960 and Middlebrook 7H10 media to encompass low-level resistance.[9]

Despite the large RMR-TB burden globally, little is known about the emergence and evolution of RMR-TB compared to MDR-TB. In addition, while the prevalence of discordant or low-level *rpoB* variants likely varies by setting [10-12], association with varying prevalence of RMR-TB is unknown. Given the high and increasing prevalence of RMR-TB in South Africa, we aimed to describe RMR-TB in detail in Khayelitsha, a peri-urban district in Cape Town, South Africa. This included risk factors for RMR-TB, the distribution of RR-conferring mutations determined through whole genome sequencing (WGS), and RIF MICs among a subset of isolates displaying *rpoB* mutations described as conferring low-level RIF resistance.

## Methods

This retrospective, cross-sectional study received ethical approval from both the University of Cape Town (UCT HREC 416/2014) and Stellenbosch University (SU N09/11/296). Patient consent for storage and sequencing of TB isolates was waived.

### Study setting and routine RR-TB diagnosis

Khayelitsha has an estimated population of 450,000 individuals with high levels of unemployment and poverty. The annual RR-TB case notification rate is estimated at 55/100,000/year and approximately 70% of RR-TB patients are HIV-positive.[13] Since 2008, most RR-TB patients are managed as outpatients with clinical, demographic and routine laboratory data collected routinely as previously described.[13]

In late 2011, Xpert MTB/RIF was introduced for routine diagnosis of TB including detection of RR among all individuals with presumptive TB; prior to this, only high-risk individuals, such as those with previous TB treatment, were tested for RR-TB. Mycobacterial culture is routinely done on samples from HIV-positive patients with presumptive TB, in whom Xpert MTB/RIF is negative for TB diagnosis, and on samples from patients with RR-TB. Line probe assay (LPA) testing is subsequently done to confirm RR and determine INH resistance on all RR-TB isolates. Once RR is diagnosed, either with Xpert MTB/RIF (or more recently Xpert MTB/RIF Ultra) or with LPA, second-line TB drug resistance testing is done. Specimens from patients with RR-TB but INH susceptibility on LPA testing, are further tested for phenotypic INH resistance at a CC of 0.1µg/ml.

### Whole genome sequencing

Individual, patient-level clinical data from RR-TB patients diagnosed between 2008 and 2017 were linked to RR-TB isolates routinely stored at -80°C in a biobank. Matched, stored isolates closest to the date of first RR-TB diagnosis were sub-cultured into *M*.*tb* BACTEC Mycobacteria Growth Indicator Tubes (MGITs) for subsequent DNA extraction and quantitative phenotypic DST (q pDST).

Genomic DNA was extracted using the phenol-chloroform method as previously described.[14] DNA concentrations were measured using Nanodrop ND-1000 spectrophotometer and DNA integrity was checked by agarose gel electrophoresis (1% gel). WGS was performed on libraries prepared from purified genomic DNA using Illumina Nextera ^®^ XT library and NEBNext ^®^ Ultra TM II FS DNA Library Prep Kits. Sequencing was performed using the Illumina HiSeq 2500 or NextSeq 500 platforms. WGS based drug resistance profiles and RR-conferring mutations were determined using TB Profiler (command line, version 2.8.12).[15] WGS data were excluded if the mean read depth across drug resistance conferring sites was <20. The *M*.*tb* numbering system was used to describe *rpoB* mutations.[16]

### Semi-quantitative phenotypic drug susceptibility testing

Based on WGS data, a convenience sample of RR-TB isolates (including MDR-TB and RMR-TB) identified with a range of common minimal or moderate confidence RR-conferring mutations[17] were tested for MIC determination. RIF MICs were determined using the BACTEC MGIT 960 system, as recommended by the manufacturer (BACTEC MGIT, Becton Dickinson, MD, USA) at doubling drug concentrations ranging from 0.03 to 1.0 µg/ml, including 2.0, 6.0, 10 and 20 µg/ml. A fully susceptible *M*.*tb* H37Rv (ATCC 27294), strain was used for quality assurance purposes to confirm the precision of each batch of reagents and drugs.

### Data analysis

RMR-TB was defined as RIF resistance and INH susceptibility regardless of other TB drug resistance, while MDR-TB was defined as resistance to both RIF and INH, again regardless of other TB drug resistance, including second-line TB drug resistance.[1] Previous TB treatment was defined for a patient who had received ≥1 month of anti-TB drugs in the past. RR-conferring mutations were classified as minimal, moderate and high-confidence in conferring RR, as previously described.[17] Chi-squared analyses were used to compare proportions and multivariate logistic regression analyses were used to assess variables associated with RMR-TB and the presence of low-level RR-conferring *rpoB* mutations. Variables were entered into multivariate models based on univariate significance or potential relevance based on literature. Data were analysed with SPSS (IBM Statistics, version 26).

## Results

### RR-TB cohort

Between 2008 and 2017 inclusive, 2,161 individuals were diagnosed with bacteriologically confirmed RR-TB in Khayelitsha. Among these, 120 (5.6%) were excluded from the cohort as they were diagnosed with RR-TB solely on the basis of an Xpert MTB/RIF or Xpert Ultra test result, without further DST to confirm RR or diagnose INH resistance. Valid WGS sequencing data were available for 1,207/2041 (59.1%) patients; however. RR-TB was identified by TB Profiler in 1,119/1,207 (92.7%) isolates and among these, 25 underwent RIF MIC determination (Figure 1).

**Figure 1:**
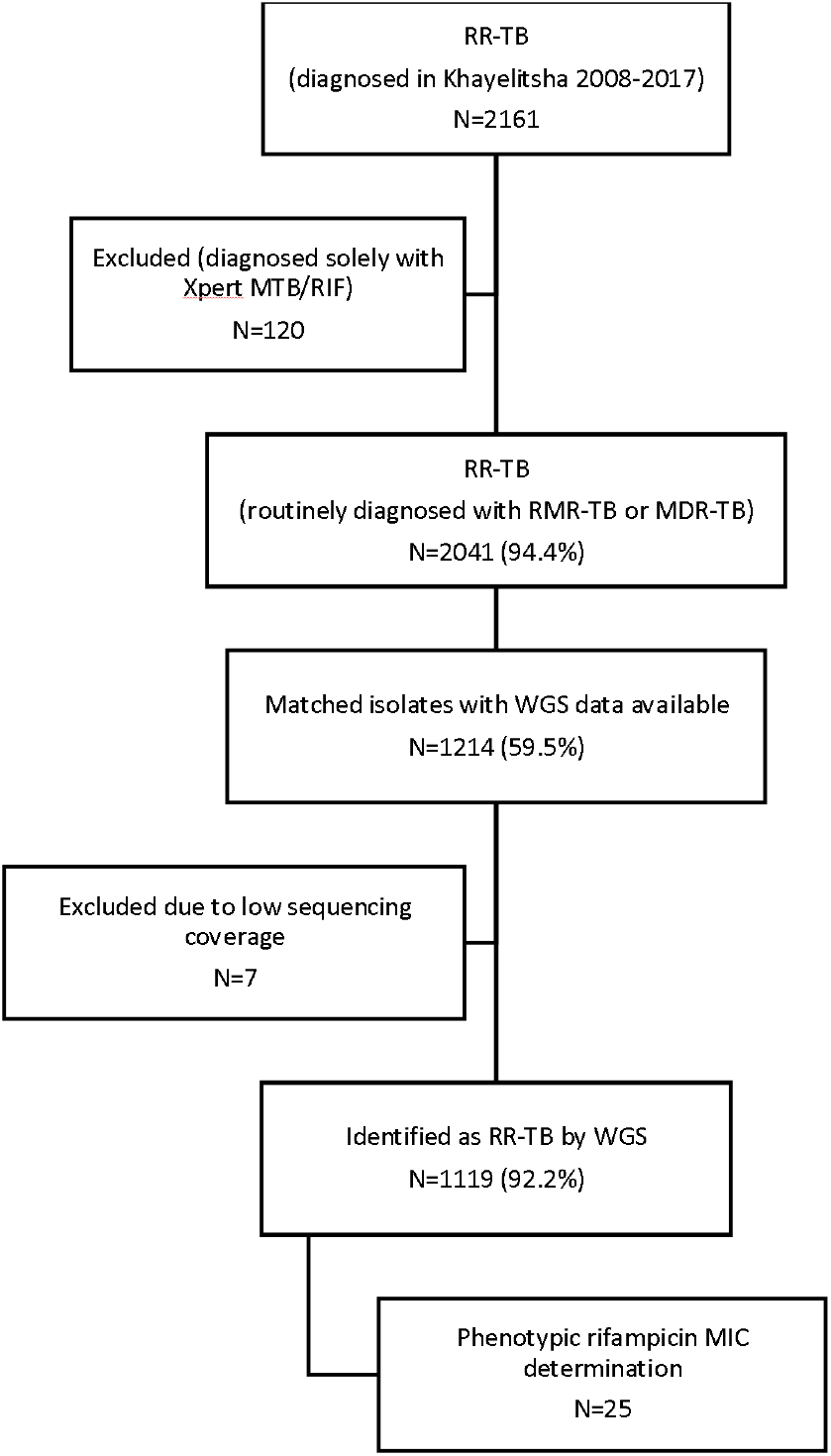
Schematic showing cohort size, availability of whole genome sequencing data and subset with MIC determination.

### Routine RMR-TB diagnosis

Overall, 463/2,041 (22.7%) individuals were diagnosed with RMR-TB. On univariate analysis, HIV-positive individuals were more likely to have RMR-TB than MDR-TB compared to those who were HIV-negative (Table 1). RMR-TB also comprised a greater proportion of all RR-TB in the second half of the study decade. On multivariate analysis, HIV-positivity, age between 35-44 years and diagnosis in the second half of the study period were significantly associated with RMR-TB compared to MDR-TB (Table 1).

**Table 1:** Association between demographic and clinical factors and routinely diagnosed RMR-TB among RR-TB patients in Khayelitsha between 2008 and 2017 inclusive.

|  | **Total**<br>**N=2,041** | **RMR-TB**<br>**N=463, N (%)** | **P value**<br>**(univariate\*)** | **Multivariate OR**<br>**(95% confidence interval)** |
| --- | --- | --- | --- | --- |
| Sex |  |  | 0.85 |  |
| Female | 991 | 223 (22.5) |  | 0.90 (0.73-1.1) |
| Male | 1050 | 240 (22.9) |  | 1.0 |
| Age (years) |  |  | 0.25 |  |
| 0-24 | 319 | 76 (23.8) |  | 1.0 |
| 25-34 | 744 | 184 (24.7) |  | 0.91 (0.66-1.3) |
| 35-44 | 634 | 131 (20.7) |  | **0.68 (0.48-0.97)** |
| 45+ | 344 | 72 (20.9) |  | 0.73 (0.50-1.1) |
| HIV status |  |  | 0.04 |  |
| Negative | 503 | 95 (18.9) |  | 1.0 |
| Positive | 1490 | 354 (23.8) |  | **1.4 (1.1-1.9)** |
| Unknown | 48 | 14 (29.2) |  | **2.5 (1.2-5.1)** |
| Previous TB treatment |  |  | 0.37 |  |
| No | 622 | 135 (21.7) |  | 1.0 |
| Yes | 1349 | 316 (23.4) |  | 1.1 (0.90-1.4) |
| Unknown | 70 | 12 (17.1) |  | 0.62 (0.31-1.2) |
| Year diagnosed |  |  | <0.01 |  |
| 2008-2012 | 1066 | 219 (20.5) |  | 1.0 |
| 2013-2017 | 975 | 244 (25.0) |  | **1.3 (1.1-1.7)** |

\*Chi-squared for difference in proportions

### Detection of rifampicin and other TB drug resistance using whole genome sequencing

WGS data were significantly more likely to be available from patients who were HIV-positive or had been previously treated for TB, although these differences were small overall (Table 2). Sequencing data were also more likely to be available when patients initiated RR-TB treatment.

**Table 2:** Comparison between patients with available TB isolate WGS data and those without.

|  | **WGS not available**<br>**N (%)** | **WGS available**<br>**N (%)** | **P value\*** |
| --- | --- | --- | --- |
| **Total N** | 827 | 1214 |  |
| Female | 416 (50.3) | 575 (47.4) | 0.21 |
| Median age (IQR) | 34 (27-41) | 34 (28-41) | 0.70 |
| HIV-positive (% of known) | 625 (75.6) | 865 (71.3) | 0.01 |
| Previous TB treatment | 535 (64.7) | 814 (67.1) | 0.01 |
| Year diagnosed (% by year; row) |  |  |  |
| 2008-2012 | 423 (39.7) | 643 (60.3) | 0.44 |
| 2013-2017 | 404 (41.4) | 571 (58.6) |  |
| RMR-TB (routine diagnosis) | 202 (24.5) | 261 (21.5) | 0.13 |
| Initiated RR-TB treatment | 679 (82.1) | 1107 (91.2) | <0.01 |

\*Chi-squared for difference in proportions

Among the 1,119 isolates where mutations known to confer RR were found, 230 (20.6%) were identified as RMR-TB and 899 (79.4%) were MDR-TB. There were clear differences in the distribution of RR-conferring mutations between RMR-TB and MDR-TB isolates (Table 3). Notably, the common high confidence *rpoB* S450L mutation was identified in only 73/230 (31.7%) RMR-TB isolates compared to 625/889 (70.3%) MDR-TB isolates (p<0.001). In contrast, the *rpoB* L430P mutation, previously described as conferring low-level RR, was identified in 32/230 (13.9%) RMR-TB isolates, compared to only 10/889 (1.1%) MDR-TB isolates (p<0.001). Overall, high confidence RR-conferring mutations were identified in 162/230 (70.4%) of RMR-TB isolates compared to 811/889 (90.2%) of MDR-TB isolates (p<0.01).

**Table 3:**
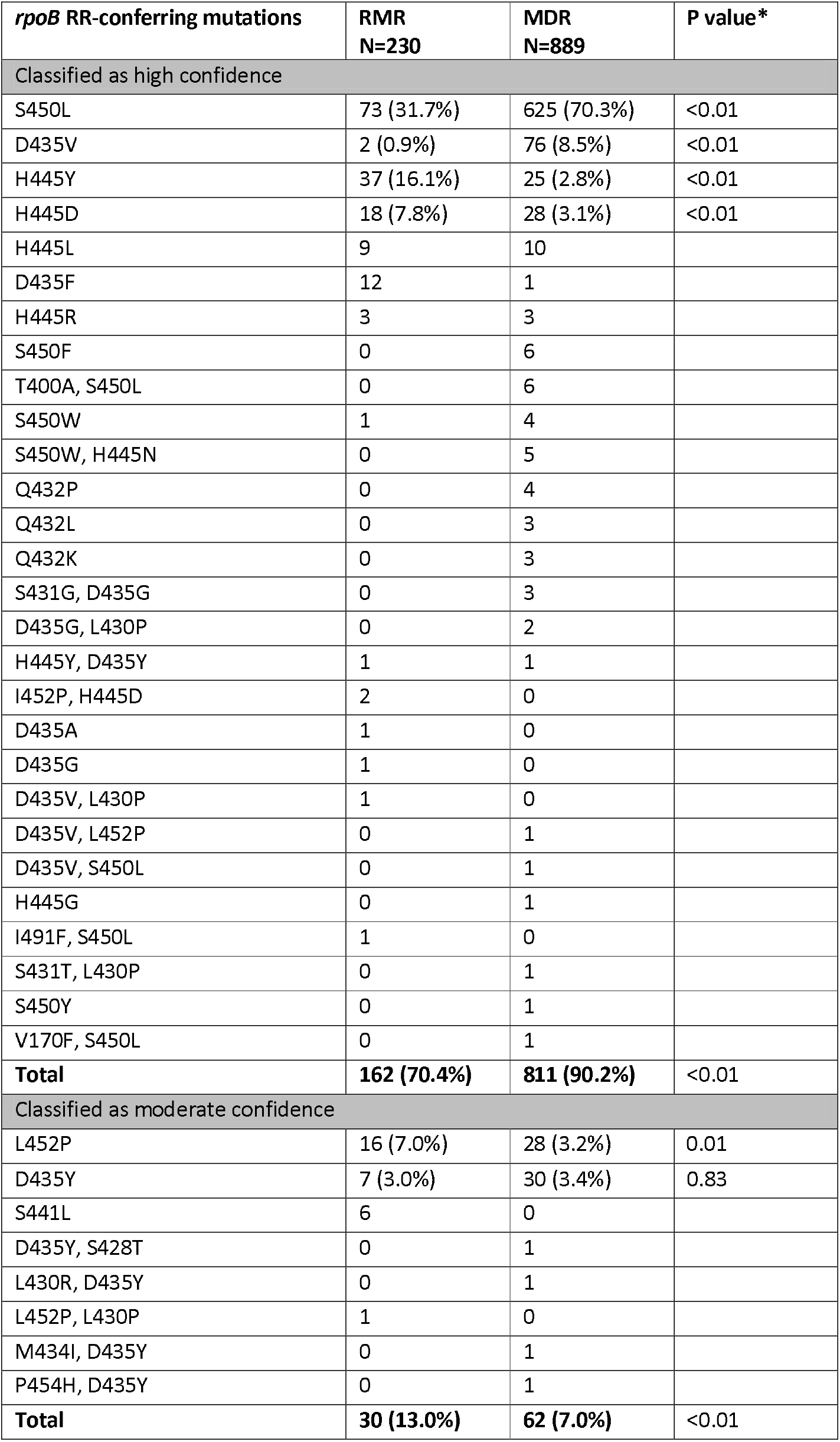
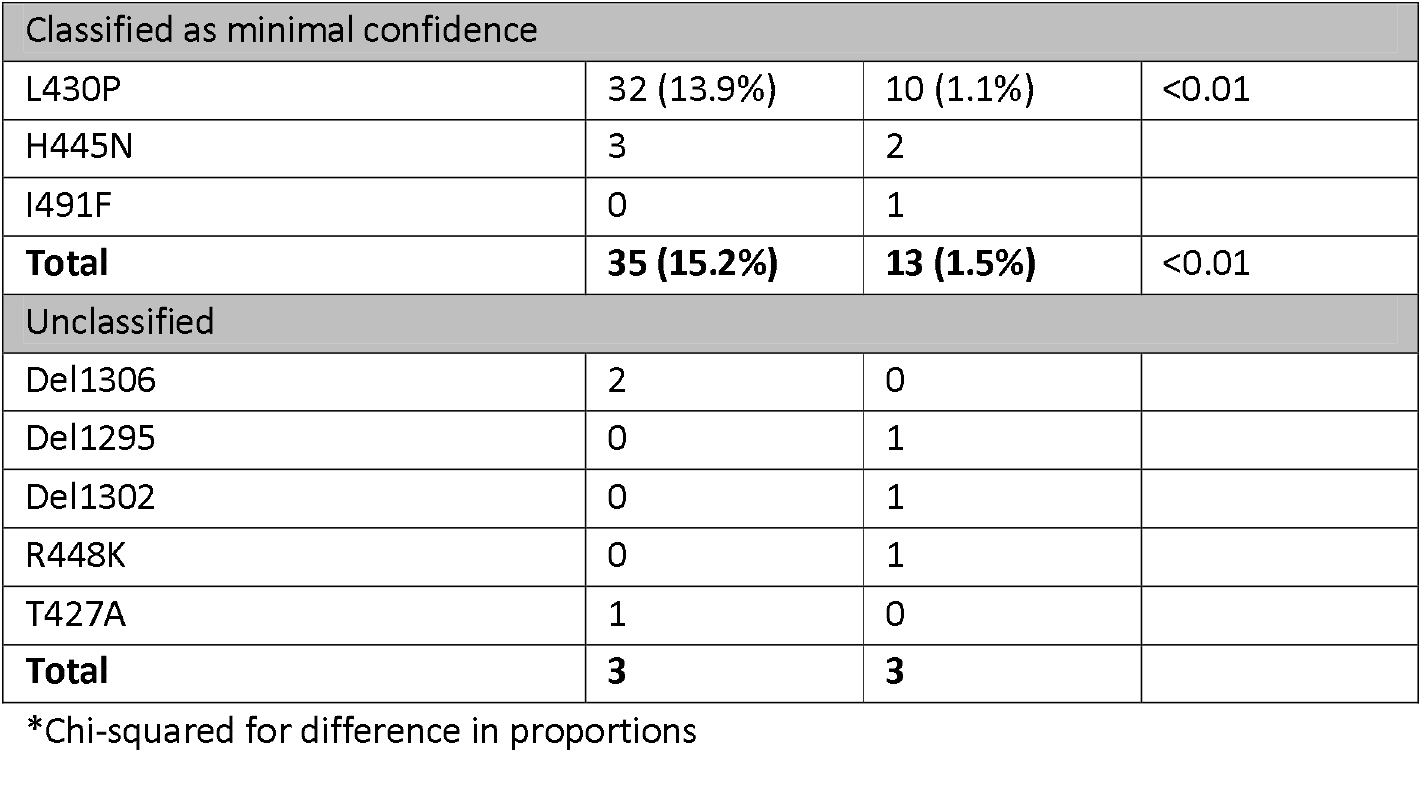
Comparison of *rpoB* mutations between RMR-TB and MDR-TB isolates and description of the confidence level for specific RR-conferring mutations (where >1 mutation was identified, the highest confidence mutation was specified).

The presence of additional TB drug resistance was also strikingly different between RMR-TB and MDR-TB isolates. Only 15/230 (6.5%) RMR-TB isolates displayed additional drug resistance conferring mutations. This contrasts with MDR-TB isolates, where 815/899 (90.7%) showed other resistance conferring mutations, in addition to those conferring RIF and INH resistance (Table 4).

**Table 4:** Complete drug resistance profile based on WGS among MDR-TB and RMR-TB isolates.

| MDR-TB |  | RMR-TB |  |
| --- | --- | --- | --- |
| Drug resistance profile | N (%) | Drug resistance profile | N (%) |
| HRZE ETH | 171 (19.2) | R | 215 (93.5) |
| HR ETH | 135 (15.2) | R ETH | 4 (1.7) |
| HR | 84 (9.4) | R INJ | 3 (1.3) |
| HRE ETH | 72 (8.1) | RZ | 3 (1.3) |
| HRE | 63 (7.1) | RE | 2 (0.9) |
| HRZE FLQ ETH | 63 (7.1) | R FLQ | 1 (0.4) |
| HRZ ETH | 61 (6.9) | RE ETH | 1 (0.4) |
| HRZE FLQ INJ ETH | 54 (6.1) | RZE | 1 (0.4) |
| HRZE INJ ETH | 46 (5.2) |  |  |
| HRZE | 42 (4.7) |  |  |
| HRZE FLQ INJ ETH CYC | 17 (1.9) |  |  |
| HRZ | 13 (1.5) |  |  |
| HRZE INJ ETH CYC | 9 (1.0) |  |  |
| HRE FLQ ETH | 8 (0.9) |  |  |
| HRE FLQ INJ ETH | 7 (0.8) |  |  |
| HRZE FLQ ETH CYC | 7 (0.8) |  |  |
| HRZ FLQ ETH | 6 (0.7) |  |  |
| HRZE ETH CYC | 5 (0.6) |  |  |
| HRZ PAS | 4 (0.4) |  |  |
| HRZE FLQ INJ | 4 (0.4) |  |  |
| HRZ INJ ETH | 3 (0.3) |  |  |
| HRZE FLQ | 3 (0.3) |  |  |
| HRE FLQ | 2 (0.2) |  |  |
| HRE INJ ETH | 2 (0.2) |  |  |
| HRZE FLQ ETH PAS | 2 (0.2) |  |  |
| HR DEL | 1 (0.1) |  |  |
| HR FLQ ETH | 1 (0.1) |  |  |
| HRE INJ | 1 (0.1) |  |  |
| HRZ FLQ INJ ETH | 1 (0.1) |  |  |
| HRZE FLQ INJ ETH PAS | 1 (0.1) |  |  |
| HRZE PAS | 1 (0.1) |  |  |
| **Total** | **889** | **230** |  |

Abbreviations: H=isoniazid; R=rifampicin; Z=pyrazinamide; E=ethambutol; ETH=ethionamide; FLQ=fluoroquinolone; INJ=second-line injectables; CYC=cycloserine; PAS=*para*-aminosalicylic acid; DEL=delamanid.

### Associations with particular rpoB mutations

Given the different *rpoB* mutation distributions, we assessed factors associated with the S450L mutation conferring high level RR and the L430P associated with low-level RR. On multivariate analysis, only MDR-TB was significantly associated with the S450L *rpoB* mutation. Similar results were seen for associations with any high confidence *rpoB* mutation (data not shown). In contrast, RMR-TB, being female and no previous TB treatment were associated with the *rpoB* L430P mutation (Table 5). HIV infection was not associated with either mutation on multivariate analysis.

**Table 5:**
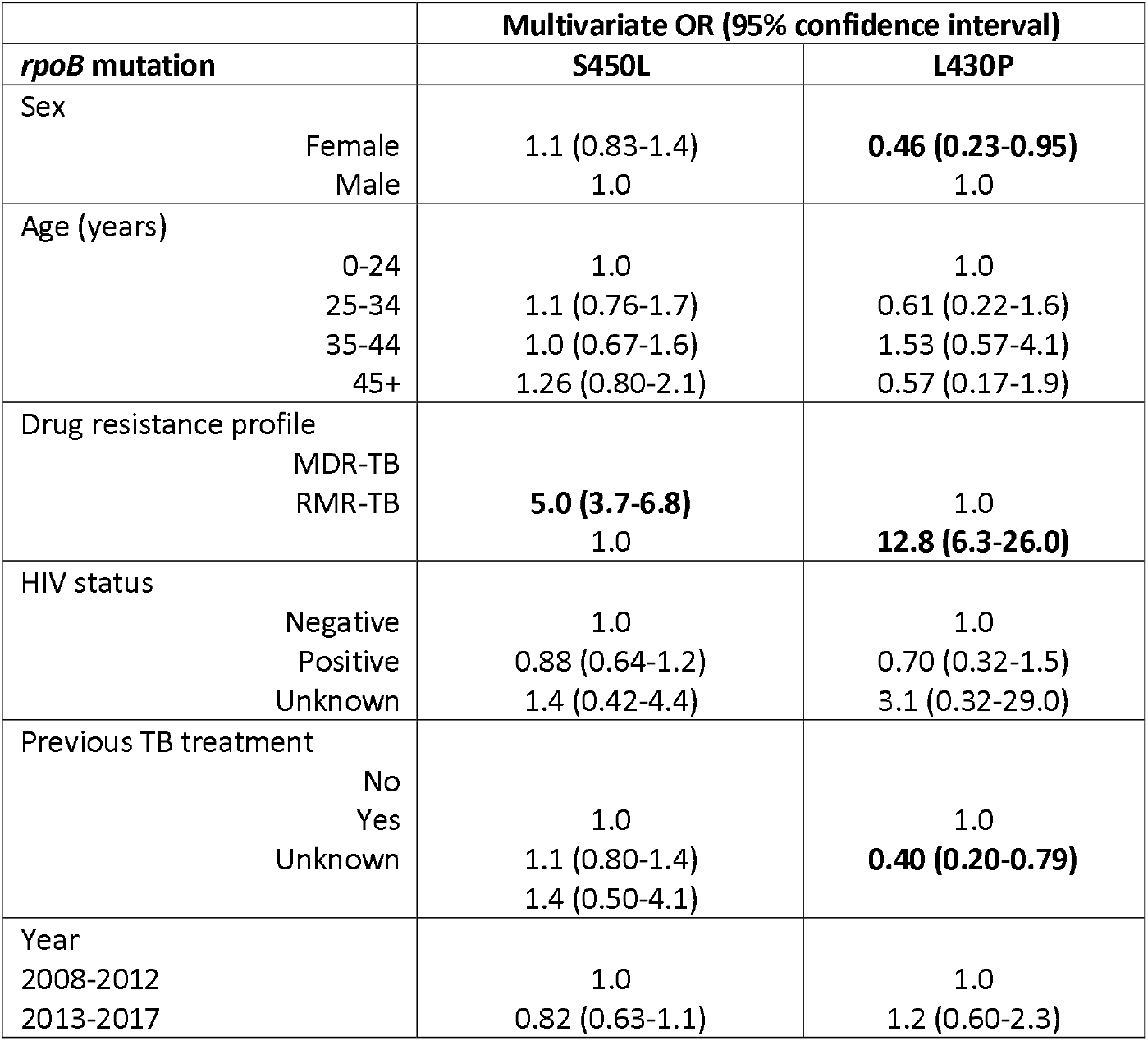
Multivariate logistic regression analysis of factors potentially associated with either the S450L or L430P *rpoB* mutations.

### Phenotypic rifampicin resistance and rpoB mutations

Quantitative phenotypic MIC testing was performed for 25 RR-TB isolates selected based on WGS data showing the most common minimal (n=13) or moderate (n=12) confidence RR-conferring mutations. Overall, 15/25 (60%) were determined to be phenotypically resistant to RIF using 0.5 µg/ml as the CC. Among the 10 isolates with the *rpoB* L430P mutation, MICs ranged from 0.125 µg/ml to 1 µg/ml, with 7 (70%) determined to be phenotypically RIF susceptible. (Table 6). Notably, all patients from whom these isolates were derived were routinely diagnosed as RR-TB with either Xpert and/or LPA.

**Table 6:**
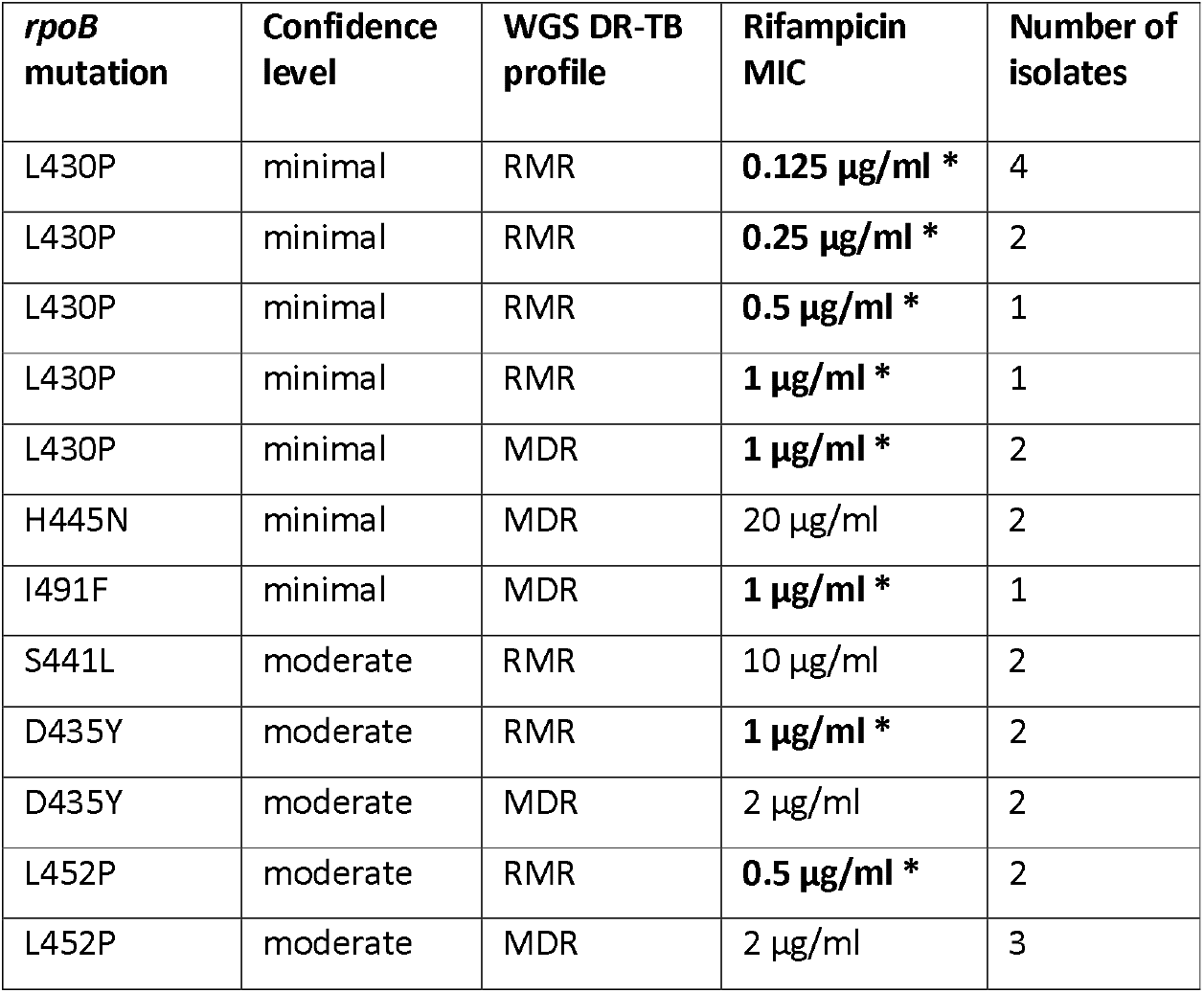
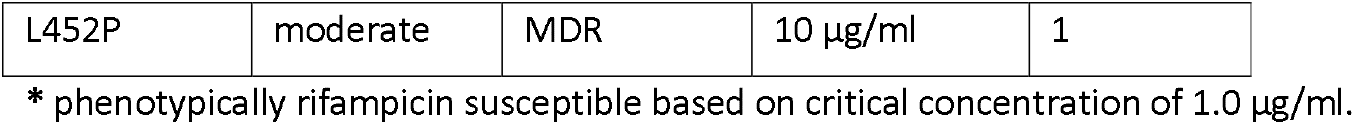
Description of quantitative phenotypic DST for rifampicin by *rpoB* mutation among 25 RR-TB isolates.

## Discussion

RMR-TB forms a significant proportion of the total RR-TB burden in this high TB, RR-TB and HIV setting. Overall, 23% of all routinely diagnosed RR-TB patients were diagnosed with RMR-TB. This figure is slightly lower than the estimate of 29% for the Western Cape Province of South Africa, and lower than the 38% reported for South Africa overall.[1, 2] There was, however, a significant increase in the proportion of RMR-TB among all RR-TB in the second half of the decade included in this study, consistent with that observed across South Africa.[2]

In this large cohort, there were significant differences in the distribution of RR-conferring mutations between RMR-TB and MDR-TB isolates. High confidence RR-conferring mutations were more commonly found among MDR-TB isolates compared to RMR-TB; only 70% of RMR-TB isolates were found to have mutations described as high confidence in conferring RIF resistance. This is similar to recent data from New York, where RMR-TB was also associated with low confidence *rpoB* mutations and low-level phenotypic RR.[18] In particular, in our setting, the most common *rpoB* S450L mutation was identified in a much higher proportion of MDR-TB isolates compared to RMR-TB, while the rarer or ‘disputed’ *rpoB* L430P mutation, with minimal or low-level confidence in conferring RR was found in 14% of RMR-TB isolates compared to only 1% of MDR-TB isolates. While the *rpoB* L430P mutation has previously been described in various settings[11, 12, 19]; it has not been reported to be associated with RMR-TB. When semi-quantitative phenotypic DST was performed on ten isolates with the L430P mutation, the majority were RIF susceptible at the revised critical concentration of 0.5 µg/ml, suggesting that a single break point for defining resistance may not be sufficient to identify low-level resistance that may well still be clinically significant.[5, 6]

RMR-TB was also significantly associated with HIV-positivity, a finding also shown in other studies.[20-23] However, there have been few representative cohort studies assessing this association in high HIV and TB burden settings. There are several mechanisms potentially underlining any association between HIV and RMR-TB. Firstly, RMR-TB isolates may be relatively less fit than their MDR-TB counterparts, thereby leading to a greater risk of infection and disease among immunocompromised HIV-positive individuals compared to HIV-negative. A recent multicentre study found that RR-TB isolates from HIV-positive patients were more likely to carry *rpoB* mutations associated with fitness costs, although there were insufficient RMR-TB cases to confirm a specific association.[24] While the higher proportion of the *rpoB* S450L mutation, which is associated with a low or no fitness cost[25] among MDR-TB isolates in our data supports this, we did not demonstrate an independent association between HIV and the presence (or absence) of the *rpoB* S450L mutation. HIV was also not an independent predictor of the *rpoB* L430P mutation, which has been associated with delayed growth in culture, suggestive of lower bacterial fitness.[26] Secondly, HIV could be associated with the emergence of RR and RMR-TB through an increased risk of resistance acquisition during TB treatment. A particular association between HIV infection and the acquisition of RR during TB treatment, predominantly among severely immunocompromised patients, has been shown.[27-29] This may be attributed to altered pharmacokinetics, potentially associated with drug malabsorption.[30] However, while HIV-positive individuals were 40% more likely to have RMR-TB in our study, there was no independent association between RMR-TB and previous TB treatment.

In addition to the different *rpoB* mutation profile seen between RMR-TB and MDR-TB isolates, there were substantially different patterns of resistance to TB drugs other than RIF and INH. Most RMR-TB isolates were only resistant to RIF with less than 3% of isolates resistant to other first-line TB drugs. These data suggest that RMR-TB treatment regimens could be tailored to include first-line TB drugs to which the isolate remains susceptible, and potentially include increased RIF doses or treatment with other rifamycins in order to overcome low-level RIF resistance.[31-33]

Currently all RR-TB patients, including those with RMR-TB are treated with predominantly second-line TB regimens, with the addition of INH in some instances.[34] This recommendation has been reiterated by the recent WHO technical expert review group.[9] While recommended second-line RR-TB regimens have improved in recent years, they remain lengthy and poorly tolerated by patients.[35] These data also highlight the potential benefits of using whole or targeted genome sequencing to individualise RR-TB treatment, particularly for RMR-TB patients, although the wide range in MICs demonstrated here suggests that associations between the presence of specific mutations and phenotypic resistance are not always clear.[36, 37]

While there were significant differences between RR-TB patients for whom WGS data were available and those not, these were small in magnitude and therefore unlikely to have had a major impact on the striking differences seen between RMR-TB and MDR-TB isolates in this dataset. Missing sequencing data was predominantly due to lack of availability of stored isolates in the biobank, in turn likely due to logistical challenges in capturing all TB isolates that are routinely diagnosed as RR-TB over such a long period. In addition, only a small subset of isolates showing *rpoB* mutations described as having minimal or moderate confidence in conferring RR underwent phenotypic MIC determination. Enlarging this subset would provide more data on the seemingly wide variability in MICs amongst isolates with the same mutation. Finally, MICs were only determined in liquid media, whereas the solid agar proportion method may have been more sensitive in detecting low-level RIF resistance.[38]

This large cohort study describing a representative community sample of RR-TB patients shows significant differences between RMR-TB and MDR-TB isolates in terms of RR-conferring *rpoB* mutations and TB drug resistance profiles. While HIV was associated with RMR-TB overall, HIV-positivity did not appear to be related to the observed differences in *rpoB* mutation distribution. Further work on this and other cohorts is required to assess the relative contributions of transmission and resistance acquisition to both RMR-TB and MDR-TB, and particularly the potential role of HIV in the increase in RMR-TB over time.

## Data Availability

Genome sequencing data from this study is available in the European Nucleotide Archive, with the accession number PRJEB45389.

## Acknowledgements

Funding for this study was provided by both a Swiss-South Africa Joint Research Award (Reference: 107799, South African National Research Foundation and the Swiss National Science Foundation) and a Wellcome Trust Fellowship (HC, reference: 099818/Z/12/Z). ZSD was funded through a PhD scholarship from the South African National Research Foundation. We would also like to acknowledge the RR-TB patients in Khayelitsha and the health care staff who provide treatment for them.

